# A mathematical model for estimating the age-specific transmissibility of a novel coronavirus

**DOI:** 10.1101/2020.03.05.20031849

**Authors:** Ze-Yu Zhao, Yuan-Zhao Zhu, Jing-Wen Xu, Qing-Qing Hu, Zhao Lei, Jia Rui, Xing-Chun Liu, Yao Wang, Meng Yang, Li Luo, Shan-Shan Yu, Jia Li, Ruo-Yun Liu, Fang Xie, Ying-Ying Su, Yi-Chen Chiang, Yan-Hua Su, Ben-Hua Zhao, Tian-Mu Chen

**Author notes:** **Correspondence: Tianmu Chen**, State Key Laboratory of Molecular Vaccinology and Molecular Diagnostics, School of Public Health, Xiamen University, 4221-117 South Xiang’an Road, Xiang’an District, Xiamen City, Fujian Province, People’s Republic of China, **Yanhua Su**, State Key Laboratory of Molecular Vaccinology and Molecular Diagnostics, School of Public Health, Xiamen University, 4221-117 South Xiang’an Road, Xiang’an District, Xiamen City, Fujian Province, People’s Republic of China, **Benhua Zhao**, State Key Laboratory of Molecular Vaccinology and Molecular Diagnostics, School of Public Health, Xiamen University, 4221-117 South Xiang’an Road, Xiang’an District, Xiamen City, Fujian Province, People’s Republic of China. These authors contributed equally to this study (Ze-Yu Zhao, Yuan-Zhao Zhu, Jing-Wen Xu).

## Abstract

**Background:** A novel coronavirus named as “SARS-CoV-2” has spread widely in many countries since December 2019, especially in China. This study aimed to quantify the age-specific transmissibility by using a mathematical model.

**Methods:** An age-specific susceptible – exposed – symptomatic – asymptomatic – recovered – seafood market (SEIARW) model was developed based on two suspected transmission routes (from market to person and person to person). The susceptible people from Wuhan City were divided into different age groups. We used the subscript *i* and *j* to represent age group 1 to 4 (*i* ≠ *j*; 1: ≤ 14 years; 2: 15-44 years; 3: 45-64 years; 4: ≥ 65 years) and 1 to 5 (*i* ≠ *j*; 1: ≤ 5 years; 2: 6-14 years; 3: 15-24 years; 4: 25-59 years; 4: ≥ 60 years), respectively. Data of reported COVID-19 cases were collected from one published literature from 26 November to 22 December, 2019 in Wuhan City, China. The age-specific transmissibility of the virus was estimated accordingly secondary attack rate (*SAR*).

**Results:** The age-specific SEIARW model fitted with the reported data well by dividing the population into four age groups (*χ*^2^ = 4.99 × 10^−6^, *P* > 0.999), and five age groups (*χ*^2^ = 4.85 × 10^−6^, *P* > 0.999). Based on the four-age-group SEIARW model, the highest transmissibility occurred from age group 2 to 3 (*SAR*_23_ = 17.56 per 10 million persons), followed by from age group 3 to 2 (*SAR*_32_ = 10.17 per 10 million persons). The lowest transmissibility occurred from age group 1 to 2 (*SAR*_12_ = 0.002 per 10 million persons). Based on the five-age-group SEIARW model, the highest transmissibility occurred from age group 4 to 5 (*SAR*_45_ = 12.40 per 10 million persons), followed by from age group 5 to 4 (*SAR*_54_ = 6.61 per 10 million persons). The lowest transmissibility occurred from age group 3 to 4 (*SAR*_34_ = 0.0002 per 10 million persons).

**Conclusions:** SARS-CoV-2 has high transmissibility among adults and elder people but low transmissibility among children and young people.

## Introduction

In December 2019, a series of cases were identified by a novel coronavirus named severe acute respiratory syndrome coronavirus 2 (SARS-CoV-2) was reported by Wuhan City, Hubei Province, China. The disease was named as coronavirus disease 2019 (COVID-19) by the World Health Organization in 11 February 2020. The patients may be related to have contacted with Huanan Seafood Wholesale Market[1], and the virus can continuously transmit from person-to-person with basic reproduction number (*R*_0_) of 2.2 (95% confidence interval [CI], 1.4 to 3.9)[2]. Another research estimated that the *R*_0_ of SARS-CoV-2 was 2.68 (95% CI, 2.47 to 2.86)[3].

Age-related transmissibility of COVID-19 has become a public health concern. The reported cases at the early stage (before 2 January 2020) of the transmission were higher than 18 years old and mainly were on 25-64 years old[1]. A study with 425 patients indicated that the median age was 59 years (range, 15 to 89) and reported different case distributions in four age groups: 0 – 14, 15 – 44, 45 – 64, and ≥ 65 years[2]. However, there is not enough epidemiological evidence to classify the age groups in transmission. In this study, we added another age classification scenario according to our previous research on influenza that people were divided into five age groups: 0 – 5, 6 – 14, 15 – 24, 25 – 59, and ≥ 60 years[4], to better understand the transmissibility of the disease for different ages.

There were several studies focusing on mathematical modelling on COVID-19[2, 3], calculating the *R*_0_ by using the serial intervals and intrinsic growth rate[2, 5, 6], or using ordinary differential equations and Markov Chain Monte Carlo methods[3]. We also developed a Bats-Hosts-Reservoir-People (BHRP) transmission network model and simplified the BHRP model as Reservoir-People (RP) transmission network model to calculate the transmissibility of SARS-CoV-2[7]. However, there is no age-specific mathematical model for quantifying the age-specific transmissibility SARS-CoV-2.

In this study, based on the RP model which we developed, we built an age-specific susceptible – exposed – symptomatic – asymptomatic – recovered – reservoir (SEIARW) model. The age-specific SEIARW model was employed to estimate the age-specific transmissibility of SARS-CoV-2 by fitting the data from 26 November to 23 December, 2019 from published literature[1].

## Methods

### Data collection

The data of COVID-19 were collected in Wuhan City from 26 November, 2019 to 23 December, 2019 from a published literature[1], including 29 COVID-19 cases with the onset date and exposure history. The data of total population which were used in our model was from Wuhan City Statistics Bureau. The data of age group proportions, birth rate and death rate in our model was obtained from Wuhan Statistical Yearbook.

### Study design

In this study, people were divided into 4 age groups based on the published literature[1], and 5 age groups based on our previous study[4]. We used the subscript *i* and *j* to represent age group 1 to 4 (*i* ≠ *j*; 1: 0 – 14 years; 2: 15 – 44 years; 3: 45 – 64 years; 4: ≥ 65 years) and 1 to 5 (*i* ≠ *j*; 1: 0 – 5 years; 2: 6-14 years; 3: 15-24 years; 4: 25-59 years; 4: ≥ 60 years), respectively.

### Age-specific transmission model

Our previous study showed that the SEIARW model could be adopted to simulate the infectious diseases transmitted from reservoir (such as water or food) to people and from person to person[8, 9]. In this study, the age-specific SEIARW model was developed based on two transmission routes (form market to person and person to person). In the model, people were divided into five compartments: susceptible (*S*), exposed (*E*), symptomatic (*I*), asymptomatic (*A*), recovered (*R*), and the seafood market were defined as reservoir (*W*). The definitions of the six compartments were shown in Table 1. The model was based on following assumptions:

**Table 1.** Variables in the age-specific SEIARW model.

| Variables | Description | Unit |
| --- | --- | --- |
| $S_i$ | Susceptible individuals density of age group $i$ | Individuals·km<sup>-2</sup> |
| $S_j$ | Susceptible individuals density of age group $j$ | Individuals·km<sup>-2</sup> |
| $E_i$ | Exposed individuals density of age group $i$ | Individuals·km<sup>-2</sup> |
| $E_j$ | Exposed individuals density of age group $j$ | Individuals·km<sup>-2</sup> |
| $I_i$ | Infectious individuals density age group $i$ | Individuals·km<sup>-2</sup> |
| $I_j$ | Infectious individuals density age group $j$ | Individuals·km<sup>-2</sup> |
| $A_i$ | Asymptomatic individuals density age group $i$ | Individuals·km<sup>-2</sup> |
| $A_j$ | Asymptomatic individuals density age group $j$ | Individuals·km<sup>-2</sup> |
| $R_i$ | Recovered individuals density age group $i$ | Individuals·km<sup>-2</sup> |
| $R_j$ | Recovered individuals density age group $j$ | Individuals·km<sup>-2</sup> |
| $W$ | Pathogen concentration in the seafood market | Viruses·mL<sup>-3</sup> |
| $N$ | Total number of population density | Individuals·km<sup>-2</sup> |

a. Susceptible individuals become infected by contact with infected/asymptomatic people;
b. SARS-CoV-2 can transmit within each age group. The transmission rate of age group *i* and *j* are *β*_*ii*_ and *β*_*jj*_ respectively.
c. SARS-CoV-2 can be transmitted between different age groups. The transmission rate from age group *i* to *j* is *β*_*ij*_ and from *j* to *i* is *β*_*ji*_.
d. Susceptible people will be infected after contact with seafood market, and the infection rate coefficient is *β*_w_;
e. All the newborn individuals which were assumed as susceptible were added into age group 1. Each age group has a natural mortality rate. We set the natural birth rate is *br*, and the natural mortality rate is *dr*;
f. The incubation period of exposed person is 1/*ω*. The exposed person will become asymptomatic people after a latent period of 1/*ω’*. We describe *p* (0 ≤ *p* ≤ 1) as the proportion of asymptomatic infection. Exposed individuals will become asymptomatic person *A* with a daily rate of *pE*, and become symptomatic person with a daily rate of (1*-p*)*E*;
g. The transmissibility of *A* is *κ* times (0 ≤ *κ* ≤ 1) of that of *I*.
h. The individuals *I* and *A* will become recovered person (*R*) after an infectious period of 1/*γ* and 1/*γ’*. A part of *I* would die due to the infection. We assumed that the case fatality rate was *f*.
i. *I* and *A* individuals can shed pathogens into *W* with the shedding rate of *μI* and *μ’A*, where *μ* and *μ’* are the shedding coefficients;
j. SARS-CoV-2 will remove in the market after a given period (the life time of the virus is 1/*ε*), and the daily decreasing rate of the pathogen is *εW*.

The flowchart of the model was shown in Figure 1. The equations of the age-specific SEIARW model were shown as follows:

**Figure 1.**
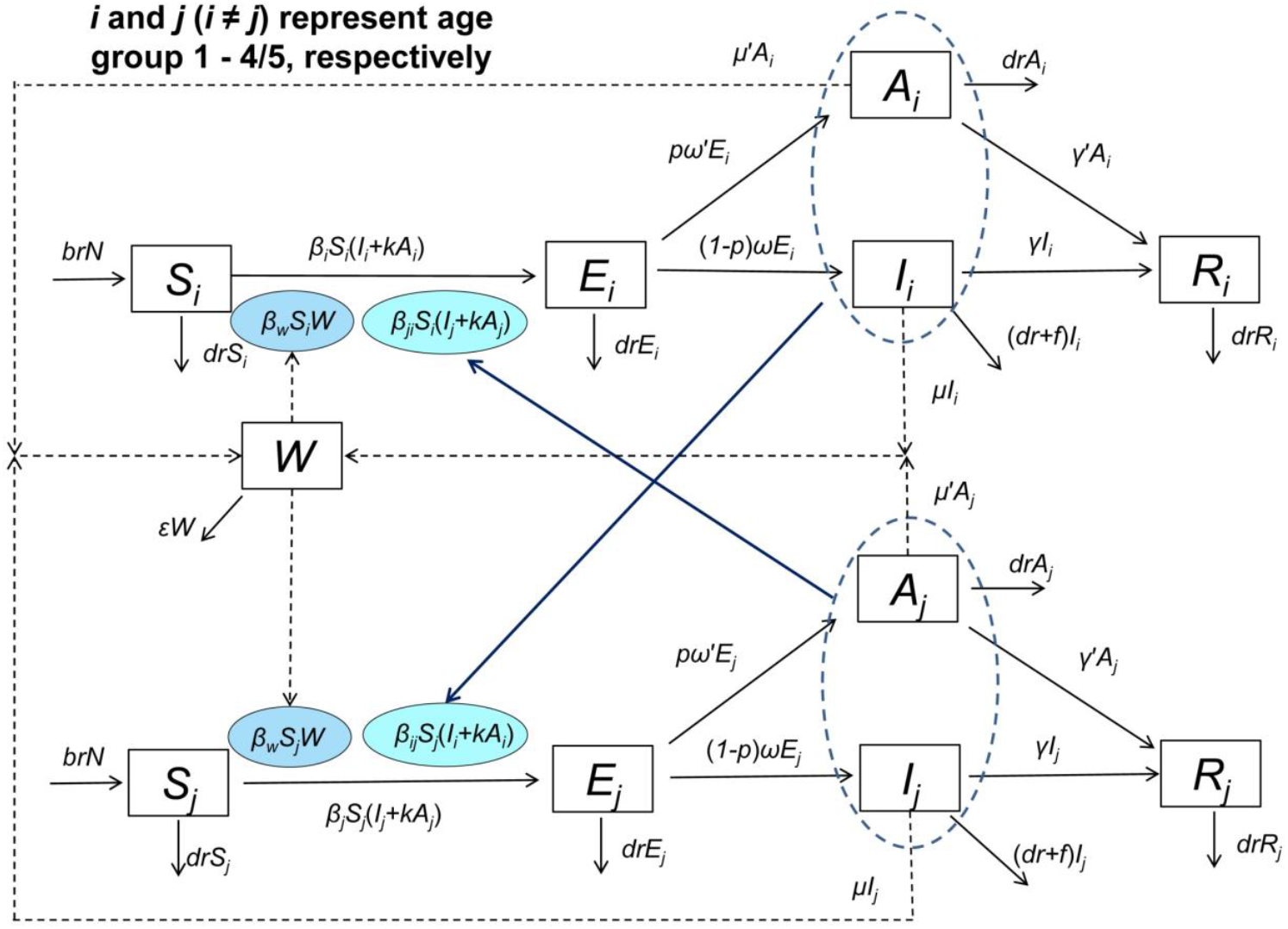
Flowchart of the age-specific SEIARW model.

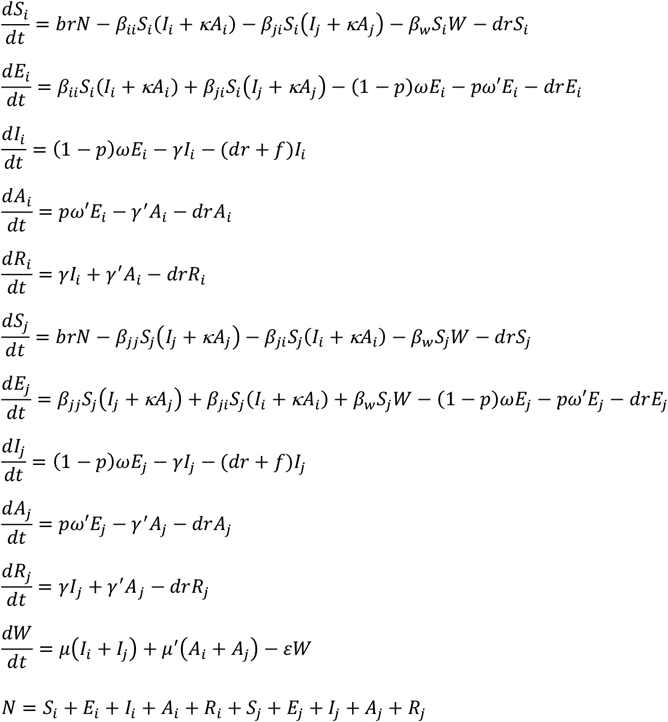

Since the different dimension of people and the virus, we adopt the following sets to perform the normalization:

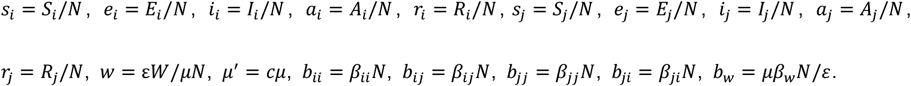

In the normalization, parameter *c* refers to relative shedding coefficients of *A* compared to *I*. The normalized model is expressed as follows:

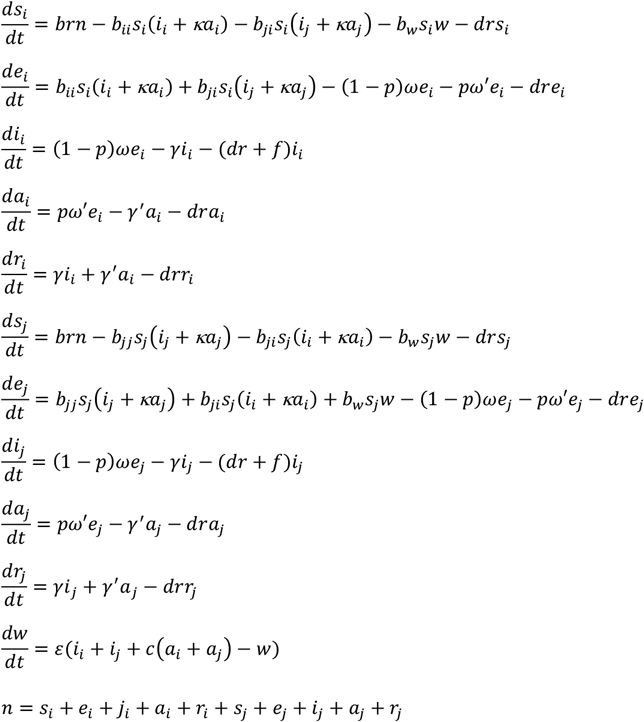

The left side of the equation shows the instantaneous rate of change of *S, E, I, A, R* and *W* at time *t*. The subscript *i* and *j* (*i* ≠ *j*) represent age group 1 to 4/5 in the equation, respectively.

### Parameters estimation

The mean incubation period was 5.2 days (95% confidence interval [*CI*]: 4.1‒7.0) [2]. As reported by Xu *et al*[10], the median time from exposure to onset of illness (infected) was 4 days (interquartile range 3-5 days). Another study showed that the mean of incubation period was around 5 days and the period falls within the range of 2-14 days [11]. We set the 5-day as the incubation period and the latent period in this study. Thus, *ω* = *ω’* = 0.2.

Since there was no data on the proportion of asymptomatic infection of the virus, we simulated the baseline value of proportion of 0.5 (*p* = 0.5). Since there was no evidence about the transmissibility of asymptomatic infection, we assumed that the transmissibility of asymptomatic infection was 0.5 times that of symptomatic infection (*κ* = 0.5), which was the similar value as influenza[12]. We assumed that the relative shedding rate of *A* compared to *I* was 0.5. Thus, *c* = 0.5. Since the SARS-CoV-2 is an RNA virus, we assumed that it could be died in the environment in a short time, but it could stay for a longer time (10 days) in the unknown hosts in the market. We set *ε* = 0.1.

There is a mean 5-day delay from symptom onset to detection/hospitalization of a case (the cases detected in Thailand and Japan were hospitalized from 3-7 days after onset, respectively) [13-15]. Another study showed that the mean time from illness onset to hospital admission (for treatment and/or isolation) was estimated at 3-4 days without truncation and at 5-9 days when right truncated[11]. As reported by Xu *et al*[10], the median time from onset of illness to first hospital admission was 2 (range from 1-4) days. A study including 45 patients diagnosed before January 1 was estimated to have a mean of 5.8 days (95% *CI*: 4.3–7.5) from illness onset to first medical visit[2]. In our model, we set the infectious period of the cases as 6 days. Therefore, *γ* = *γ’* = 0.1667.

According the official report by National Health Commission of the People’s Republic of China, we collected the data of daily fatality from January 24^th^ to 30^th^, 2020[16]. Therefore, we set *f* as 0.02348 (range: 0.02198-0.03186). In this study, we set the total population as 11,081,000 so that *br* = 4.266 ×10^−5^ and *dr* = 3.184 × 10^−5^ based on the 2018 Wuhan Statistical Yearbook (Table 2).

**Table 2.**
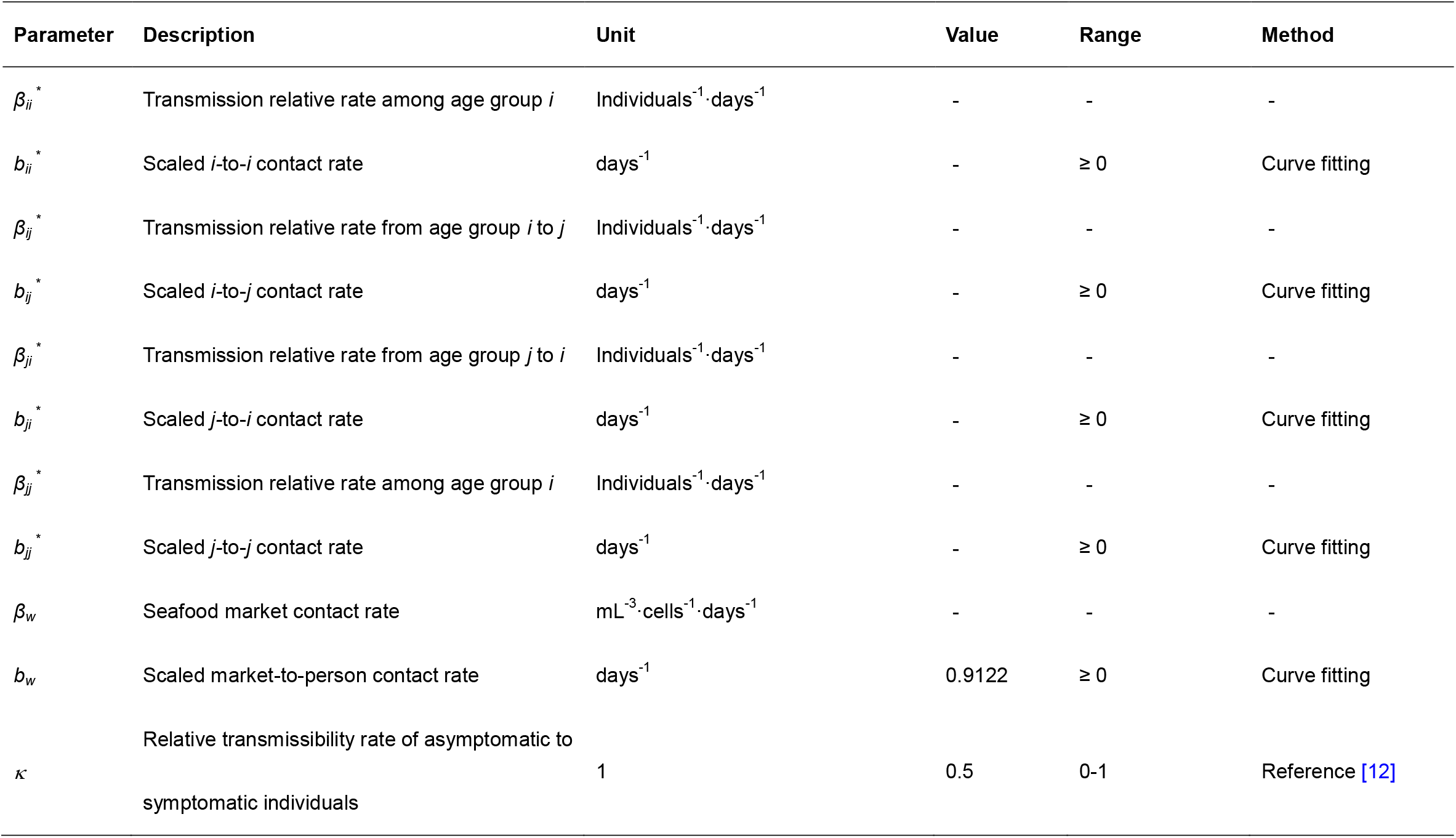
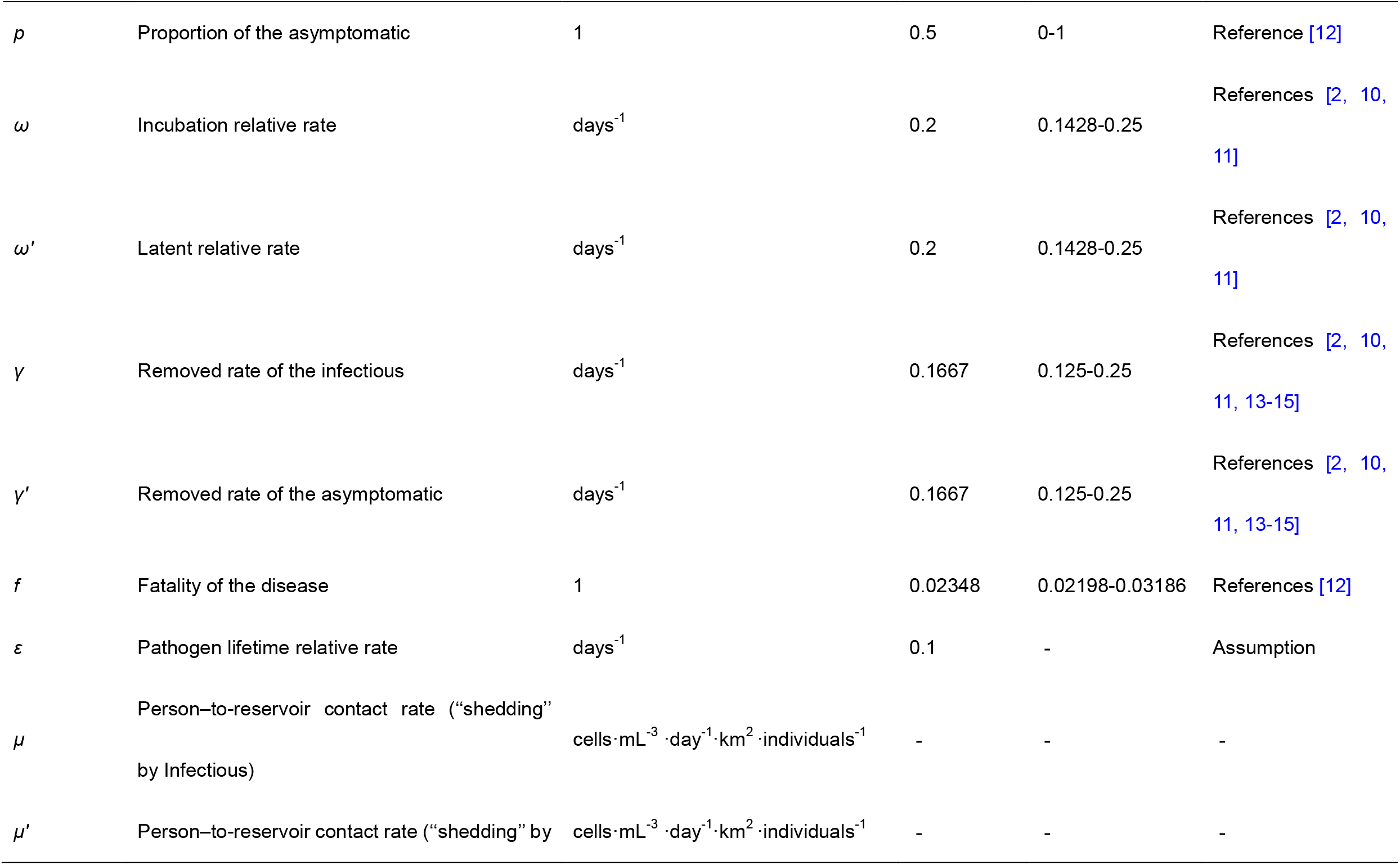
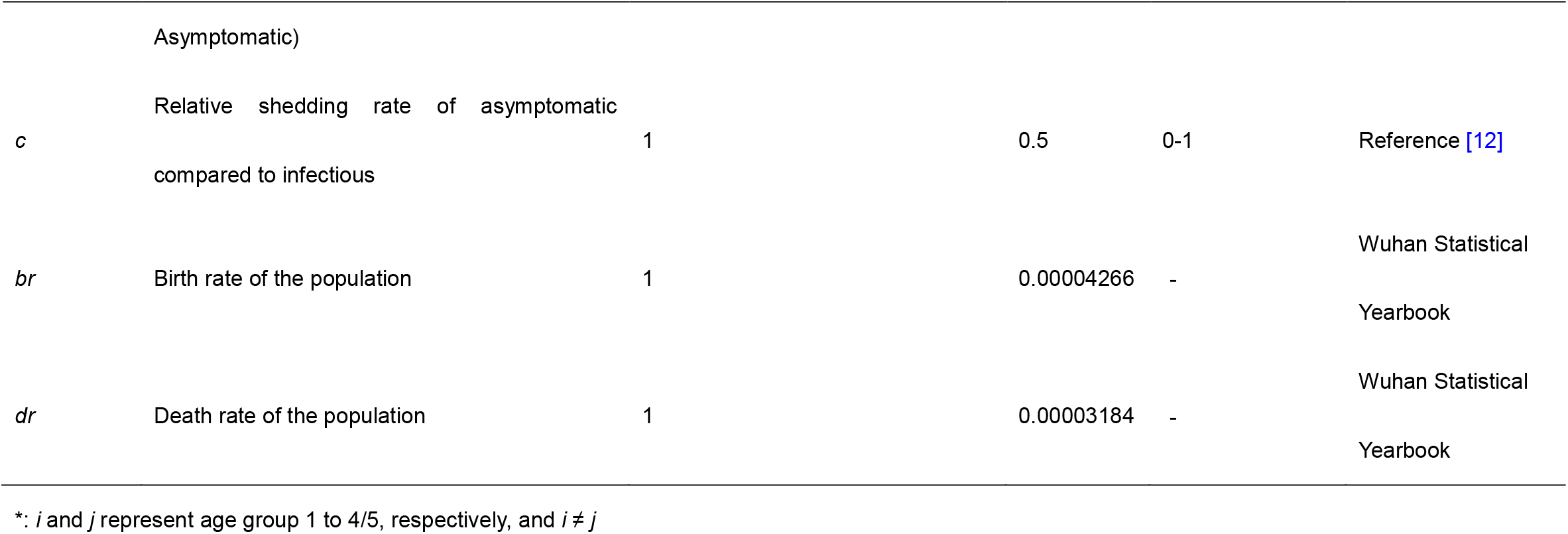
Description and values of parameters in the age-specific SEIARW model.

| Parameter | Description | Unit | Value | Range | Method |
| --- | --- | --- | --- | --- | --- |
| $\beta_{ii}^*$ | Transmission relative rate among age group $i$ | Individuals<sup>-1</sup>·days<sup>-1</sup> | - | - | - |
| $b_{ii}^*$ | Scaled $i$-to-$i$ contact rate | days<sup>-1</sup> | - | $\geq 0$ | Curve fitting |
| $\beta_{ij}^*$ | Transmission relative rate from age group $i$ to $j$ | Individuals<sup>-1</sup>·days<sup>-1</sup> | - | - | - |
| $b_{ij}^*$ | Scaled $i$-to-$j$ contact rate | days<sup>-1</sup> | - | $\geq 0$ | Curve fitting |
| $\beta_{ji}^*$ | Transmission relative rate from age group $j$ to $i$ | Individuals<sup>-1</sup>·days<sup>-1</sup> | - | - | - |
| $b_{ji}^*$ | Scaled $j$-to-$i$ contact rate | days<sup>-1</sup> | - | $\geq 0$ | Curve fitting |
| $\beta_{jj}^*$ | Transmission relative rate among age group $j$ | Individuals<sup>-1</sup>·days<sup>-1</sup> | - | - | - |
| $b_{jj}^*$ | Scaled $j$-to-$j$ contact rate | days<sup>-1</sup> | - | $\geq 0$ | Curve fitting |
| $\beta_w$ | Seafood market contact rate | mL<sup>-3</sup>·cells<sup>-1</sup>·days<sup>-1</sup> | - | - | - |
| $b_w$ | Scaled market-to-person contact rate | days<sup>-1</sup> | 0.9122 | $\geq 0$ | Curve fitting |
| $\kappa$ | Relative transmissibility rate of asymptomatic to symptomatic individuals | 1 | 0.5 | 0-1 | Reference [12] |
| $\rho$ | Proportion of the asymptomatic | 1 | 0.5 | 0-1 | Reference [12] |
| $\omega$ | Incubation relative rate | days<sup>-1</sup> | 0.2 | 0.1428-0.25 | References [2, 10, 11] |
| $\omega'$ | Latent relative rate | days<sup>-1</sup> | 0.2 | 0.1428-0.25 | References [2, 10, 11] |
| $\gamma$ | Removed rate of the infectious | days<sup>-1</sup> | 0.1667 | 0.125-0.25 | References [2, 10, 11, 13-15] |
| $\gamma'$ | Removed rate of the asymptomatic | days<sup>-1</sup> | 0.1667 | 0.125-0.25 | References [2, 10, 11, 13-15] |
| $f$ | Fatality of the disease | 1 | 0.02348 | 0.02198-0.03186 | References [12] |
| $\varepsilon$ | Pathogen lifetime relative rate | days<sup>-1</sup> | 0.1 | - | Assumption |
| $\mu$ | Person-to-reservoir contact rate ("shedding" by Infectious) | cells·mL<sup>-3</sup>·day<sup>-1</sup>·km<sup>2</sup>·individuals<sup>-1</sup> | - | - | - |
| $\mu'$ | Person-to-reservoir contact rate ("shedding" by | cells·mL<sup>-3</sup>·day<sup>-1</sup>·km<sup>2</sup>·individuals<sup>-1</sup> | - | - | - |
|  | Asymptomatic) |  |  |  |  |
| *c* | Relative shedding rate of asymptomatic compared to infectious | 1 | 0.5 | 0-1 | Reference [12] |
| *br* | Birth rate of the population | 1 | 0.00004266 | - | Wuhan Statistical Yearbook |
| *dr* | Death rate of the population | 1 | 0.00003184 | - | Wuhan Statistical Yearbook |

\*: $i$ and $j$ represent age group 1 to 4/5, respectively, and $i \neq j$

A two-step curve fitting method was adopted to estimate the parameter *b*_*W*_, *b*_*ii*_, *b*_*jj*_, *b*_*ij*_, and *b*_*ji*_. At the first step, we used the mix-age SEIARW model to fit the data and to estimate the parameter *b*_*W*_ and mix-age *b* (named as *b*_*p*_). At the second step, we used the age-specific SEIARW model to fit the data and to estimate the parameter *b*_*ii*_, *b*_*jj*_, *b*_*ij*_, and *b*_*ji*_.

### Quantification of the age-specific transmissibility of SARS-CoV-2

In the model the age-specific secondary attack rate (*SAR*) was employed to quantify the transmissibility of SARS-CoV-2. They were calculated as follows:

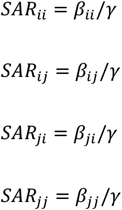

In the equations, *SAR*_*ii*_, *SAR*_*ij*_, *SAR*_*ji*_, and *SAR*_*jj*_ refer to the age-specific *SAR* among age group *i*, from age group *i* to *j*, from age group *j* to *i*, and among age group *j*, respectively.

To quantify the age-specific transmissibility of SARS-CoV-2, we also performed a simulation which was named as “knock-out” simulation in our previous study[17]. In this study, the “knock-out” simulation was defined as cutting off the route of transmission between or within different age groups, and was performed in the following scenarios: A) *β*_*ii*_ = 0; B) *β*_*ji*_ = 0; C) *β*_*ij*_ = 0; D) *β*_*jj*_ = 0; E) control (no cutting off transmission route).

### Simulation method and statistical analysis

Berkeley Madonna 8.3.18 (developed by Robert Macey and George Oster of the University of California at Berkeley. Copyright ©1993-2001 Robert I. Macey & George F. Oster) was employed to perform the curve fitting and the simulation. The simulation methods (Runge-Kutta method of order four with tolerance set at 0.001) were the same as the previously published researches [18-24]. The data was analyzed and figured by using Microsoft Office Excel 2010 (Microsoft, Redmond, WA, USA) and GraphPad Prism 7.0 (GraphPad Software, La Jolla, CA). The goodness of fit was judged by coefficient of determination (*R*^2^) and Chi-square (*χ*^2^) value calculated by SPSS 21.0 (IBM Corp, Armonk, NY, USA).

### Sensitivity analysis

Since nine parameters (*p, κ, c, ε, ω, ω’, γ, γ’* and *f*) in the models were collected from literatures, there might be some uncertainties in our model. Therefore, we performed sensitivity analysis of parameters *p, κ, c, ε, ω, ω’, γ, γ’* and *f* by splitting them into 1,000 values ranging from 0 – 0.9, 0 – 1, 0 – 1, 0 – 1, 0.1428 – 0.25, 0.1428 – 0.25, 0.125 – 0.25, 0.125 – 0.25, 0.02198 – 0.03186, respectively (Table 2).

## Results

### Epidemiological characteristics and curve fitting of SARS-CoV-2

There were 29 COVID-19 cases data collected in Wuhan City from 26 November, 2019 to 23 December, 2019, among which 10 cases had a history of the seafood market exposure and 19 cases had no history of the exposure (Figure 2).

**Figure 2.**
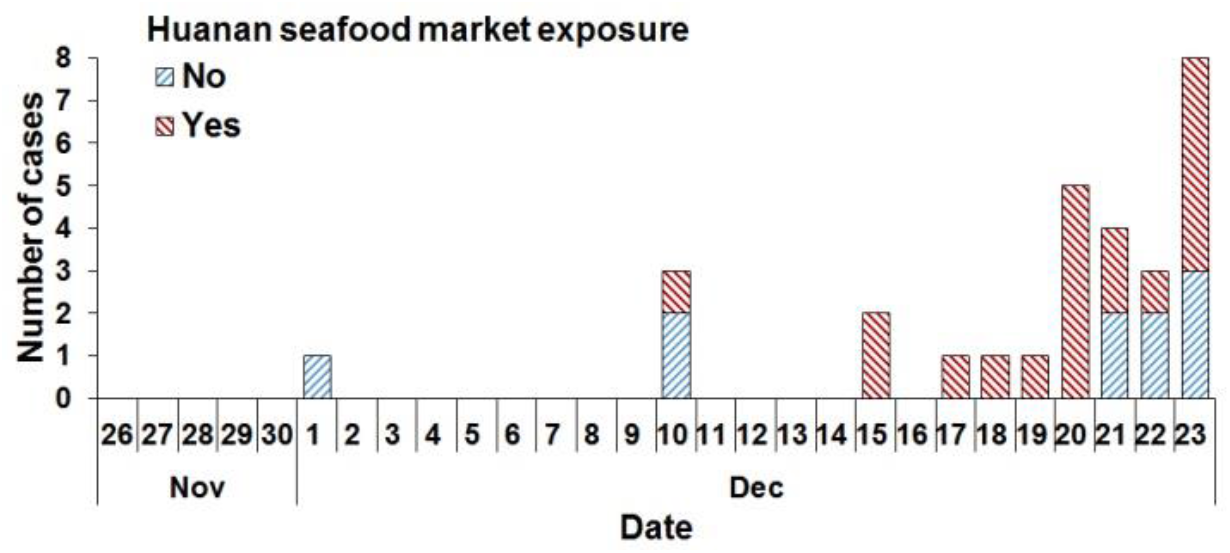
Epidemic curve of collected data of reported COVID-19 cases in Wuhan City from 26 November, 2019 to 23 December, 2019.

The mix-age SEIARW model including all age groups fitted the reported data well (*χ*^2^ = 4.77 × 10^−6^, *P* > 0.999). The results of the curve fitting were shown in Figure 3. The results also showed that the *b*_*p*_= 1.1329 and *b*_*W*_ = 0.5255. The age-specific SEIARW model fitted with the reported data well by dividing the population into four age groups (*χ*^2^ = 4.99 × 10^−6^, *P* > 0.999), and five age groups (*χ*^2^ = 4.85 × 10^−6^, *P* > 0.999). Thus, the prevalence of COVID-19 in each age group was simulated in Figure 4.

**Figure 3.**
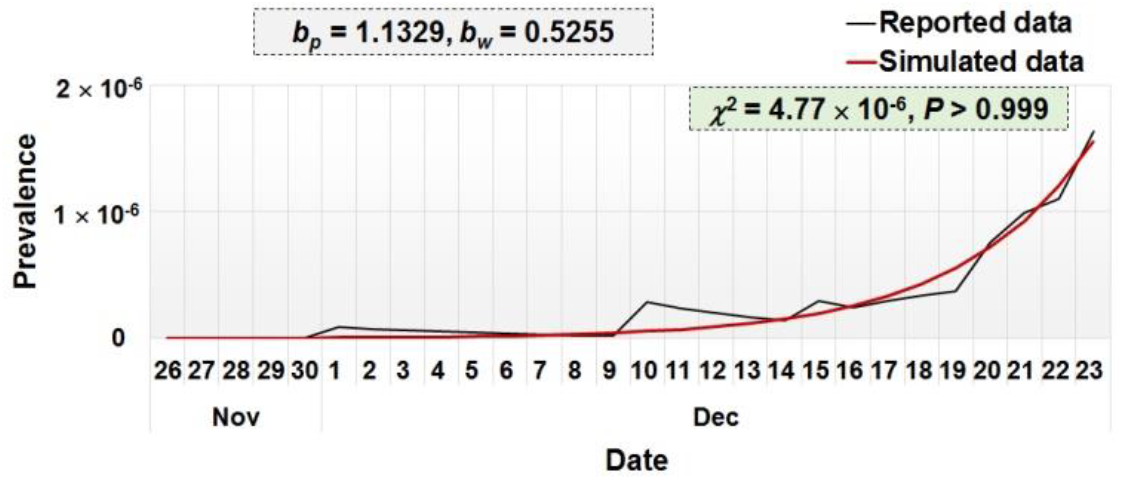
Results of curve fitting of the mix age SEIARW model to the reported data.

**Figure 4.**
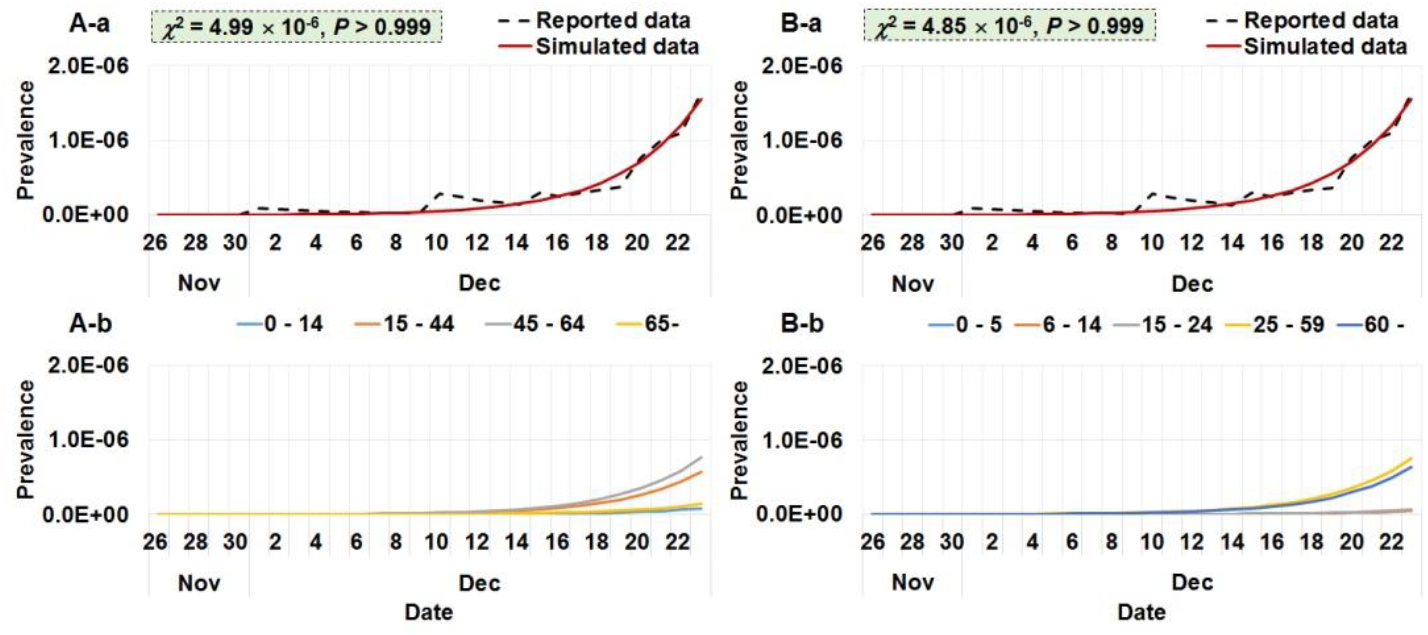
Results of curve fitting of the age-specific SEIARW model to the reported data. A-a: curve fitting based on four age groups; A-b: the simulated prevalence by the age-specific SEIARW model based on the four age groups; B-a: curve fitting based on five age groups; B-b: the simulated prevalence by the age-specific SEIARW model based on the five age groups.

### Transmissibility of SARS-CoV-2

Based on the four-age-group SEIARW model, the highest transmissibility occurred from age group 2 to 3 (*SAR*_23_ = 17.56 per 10 million persons), followed by from age group 3 to 2 (*SAR*_32_ = 10.17 per 10 million persons), from age group 4 to 4 (*SAR*_44_ = 6.99 per 10 million persons), and from age group 3 to 4 (*SAR*_34_ = 5.69 per 10 million persons). The lowest transmissibility occurred from age group 1 to 2 (*SAR*_12_ = 0.002 per 10 million persons), followed by from age group 3 to 1 (*SAR*_31_ = 0.003 per 10 million persons), from age group 4 to 2 (*SAR*_42_ = 0.52 per 10 million persons), and from age group 1 to 3 (*SAR*_13_ = 1.08 per 10 million persons) (Figure 5-A).

**Figure 5.**
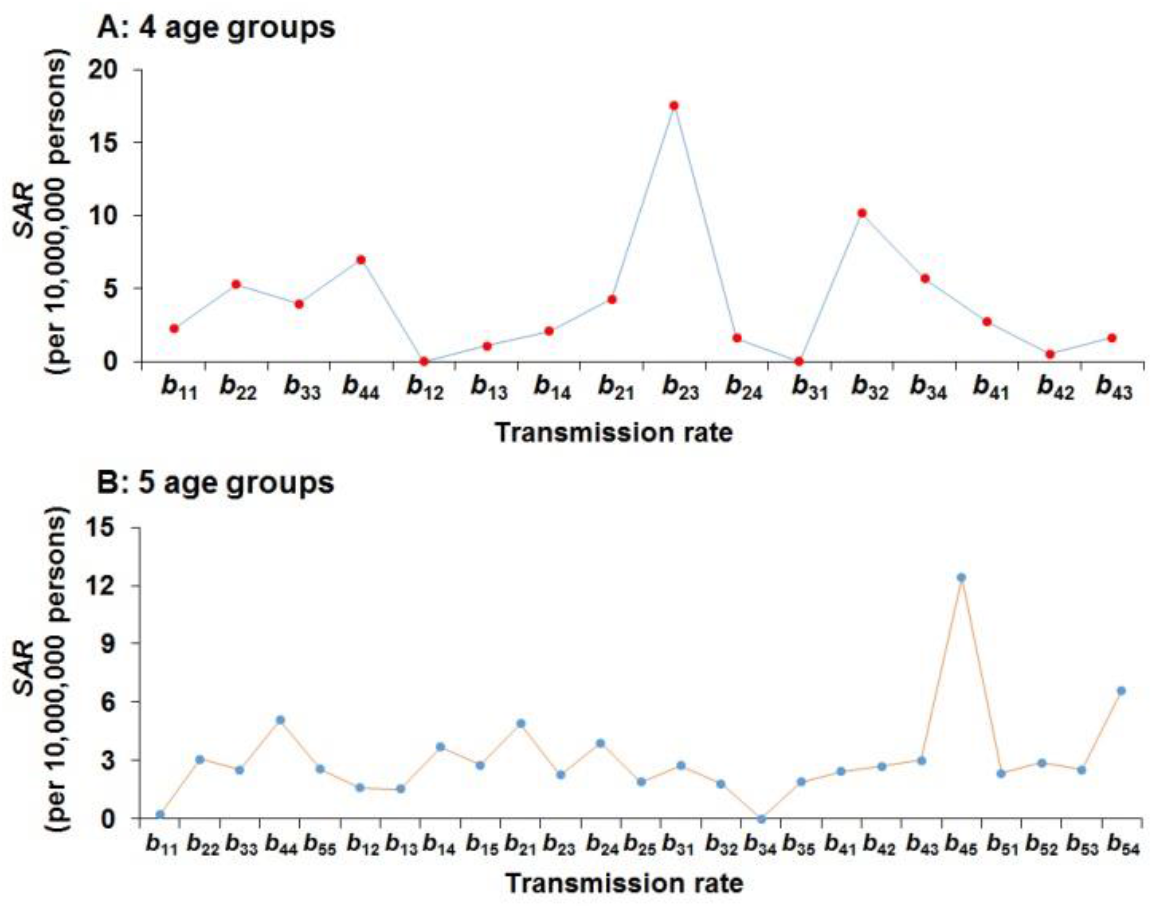
SAR simulated from the age-specific SEIARW model. A: SAR based on four age groups; B: SAR based on five age groups.

Based on the five-age-group SEIARW model, the highest transmissibility occurred from age group 4 to 5 (*SAR*_45_ = 12.40 per 10 million persons), followed by from age group 5 to 4 (*SAR*_54_ = 6.61 per 10 million persons), from age group 4 to 4 (*SAR*_44_ = 5.08 per 10 million persons), and from age group 2 to 1 (*SAR*_21_ = 4.90 per 10 million persons). The lowest transmissibility occurred from age group 3 to 4 (*SAR*_34_ = 0.0002 per 10 million persons), followed by from age group 1 to 1 (*SAR*_11_ = 0.22 per 10 million persons), from age group 1 to 3 (*SAR*_13_ = 1.54 per 10 million persons), and from age group 1 to 2 (*SAR*_12_ = 1.59 per 10 million persons) (Figure 5-B).

The results of the “knock-out” simulation showed that, based on the four-age-group SEIARW model, the scenarios *b*_23_ = 0, *b*_32_ = 0, and *b*_22_ = 0 led to the highest decrease for the number of cases. However, the scenarios *b*_12_ = 0, *b*_31_ = 0, and *b*_11_ = 0 led to the lowest decrease for the number of cases (Figure 6-A). Based on the five-age-group SEIARW model, the scenarios *b*_44_ = 0, *b*_45_ = 0, and *b*_54_ = 0 led to the highest decrease for the number of cases. However, the scenarios *b*_12_ = 0, *b*_31_ = 0, and *b*_11_ = 0 led to the lowest decrease for the number of cases (Figure 6-B).

**Figure 6.**
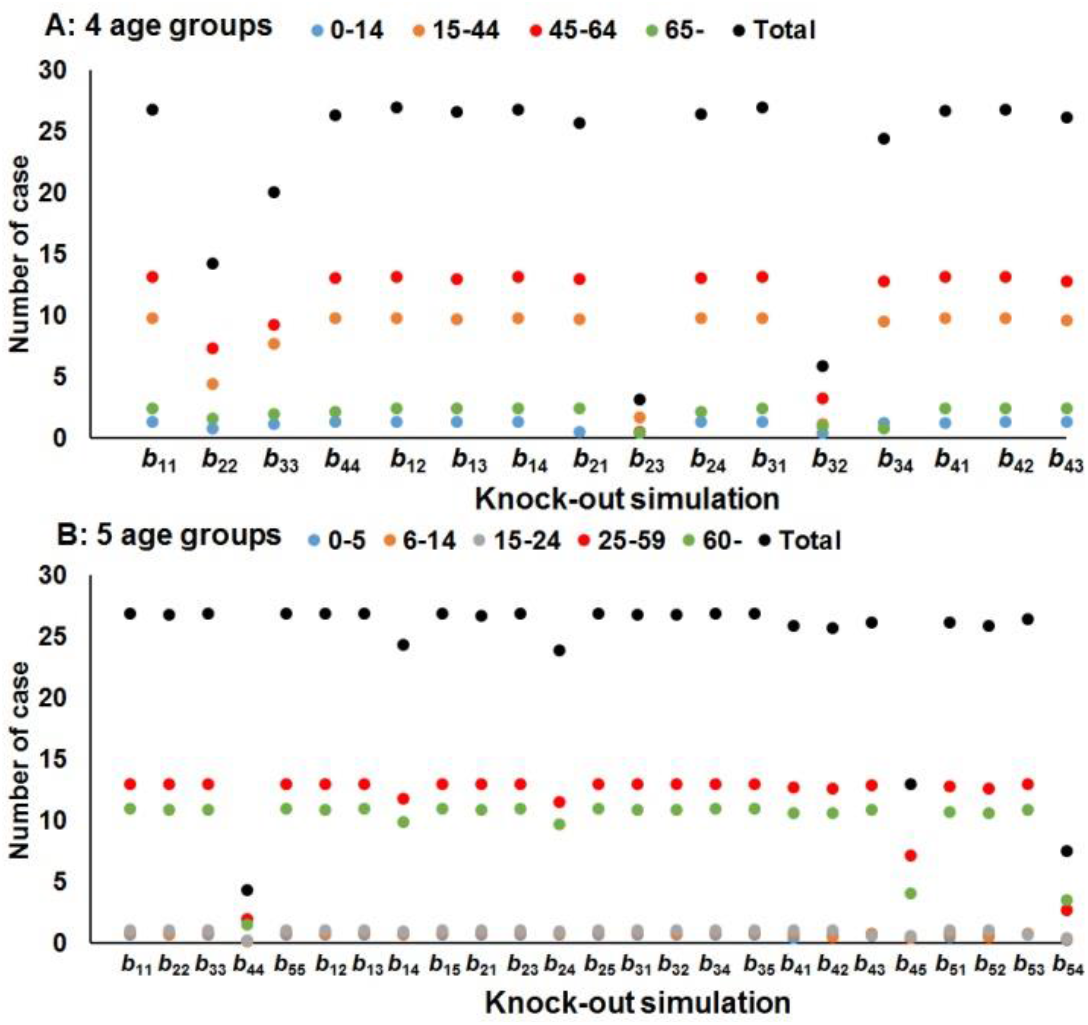
Results of the “knock-out” simulation from the age-specific SEIARW model. A: results based on four age groups; B: results based on five age groups.

### Sensitivity analysis

The results of sensitivity analysis showed that the models were slightly sensitive to parameters *p*,

*ω*, and *γ* but not sensitive to parameters *κ, c, ε, f, ω’*, and *γ’* (Figures 7 – 8).

**Figure 7.**
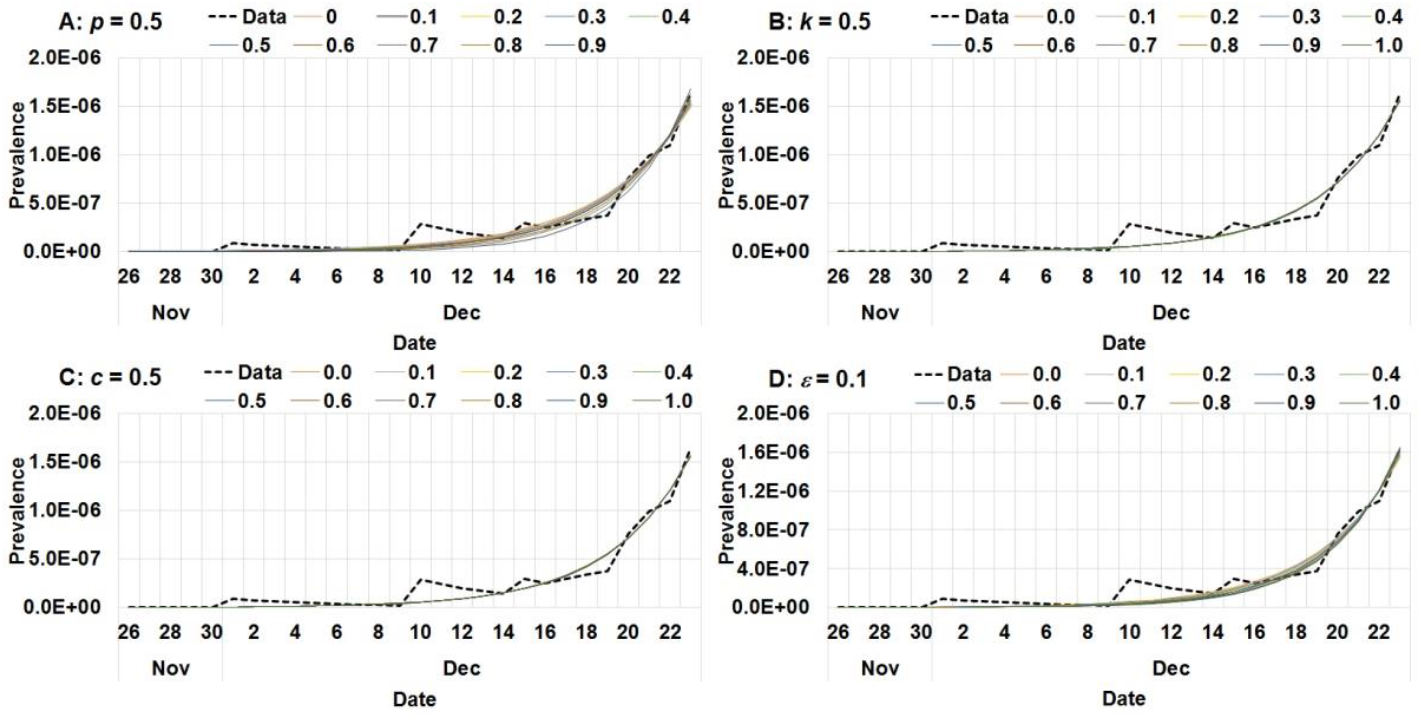
Results of sensitivity analysis of *p, κ, c*, and *ε*. A: sensitivity analysis of *p*; B: sensitivity analysis of *κ*; C: sensitivity analysis of *c*; D: sensitivity analysis of *ε*.

**Figure 8.**
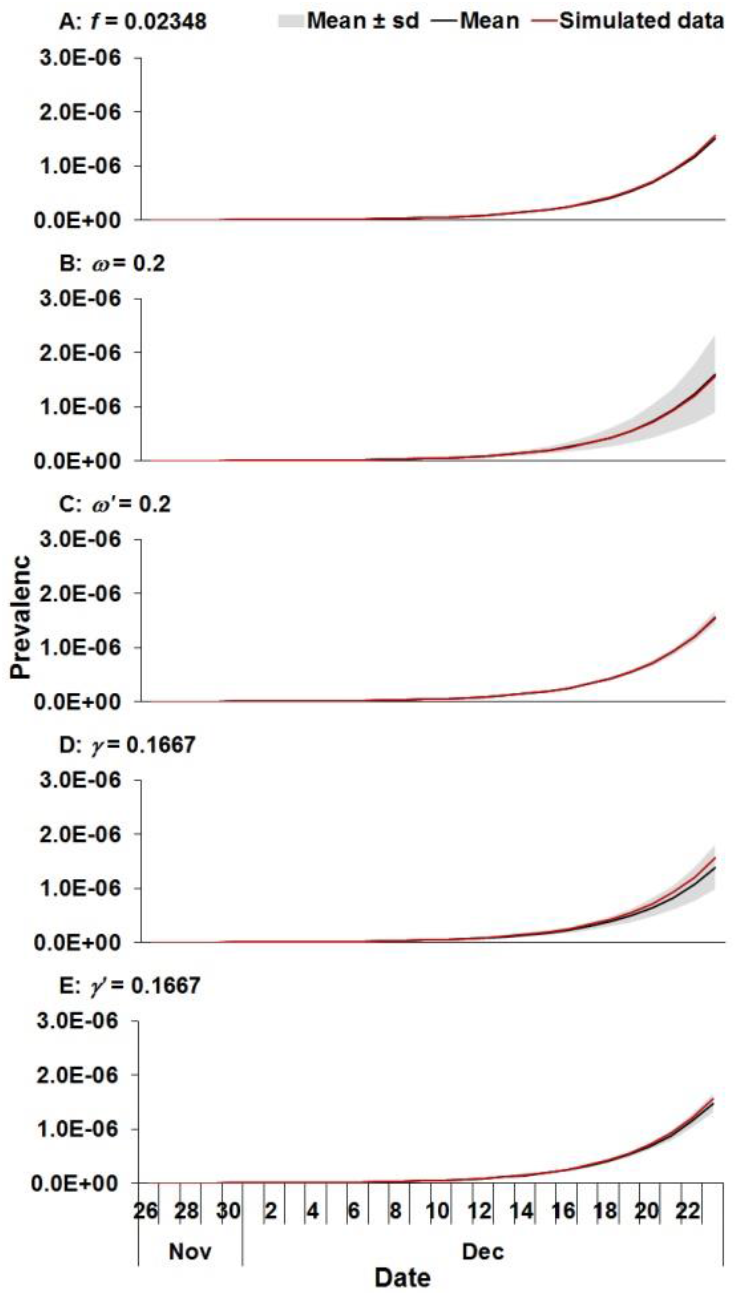
Results of sensitivity analysis of *ω, ω’, γ, γ’* and *f*. A: sensitivity analysis of *ω*; B: sensitivity analysis of *ω’*; C: sensitivity analysis of *γ*; D: sensitivity analysis of *γ’*; F: sensitivity analysis of *f*.

## Discussion

This is the first study to develop an age-specific SEIARW model to quantify the transmissibility of COVID-19 among different age groups. The results showed that our model fitted the reported data well, thus has the capability of estimating or predicting? the age-specific transmissibility of the virus.

Based on the four-age-group SEIARW model, the highest transmissibility occurred between the age groups 15 – 44 years and 45 – 64 years, among those ≥ 65 years, or from 45 – 64 years to ≥ 65 years. The lowest transmissibility occurred from age group 0-14 years to 15 – 44 years, or from 45 – 64 years to ≤14 years. Based on the five-age-group SEIARW model, the highest transmissibility occurred between age group 25 – 59 years and ≥ 60 years, or among 25 – 59 years. The lowest transmissibility occurred from age group 15 – 24 years to 25 – 59 years, or from age group 0-5 years to 6-14 years, or to 15-24 years. The “knock-out” simulation had the similar results. We concluded that the virus was more likely to transmit among older population.

These results revealed that SARS-CoV-2 has high transmissibility among adult older than 25 years old or elder people, but has low transmissibility among children or people younger than 14 years. These results were similar to the age-specific transmissibility of influenza A (H1N1)[4]. The age-specific control and prevention interventions are needed.

The reasons for the difference of age-specific transmissibility remain unclear. It might be related to the different contact characteristics among different age groups. Adults were more likely to work outside and came into contact with different individuals in work places, buses, subways, or even airplanes. However, children or younger people could stay at home during the outbreak, and they were less likely to get infected, or if they were infected, it was mostly from adults or elder people in the same family.

Our findings were based on the parameter estimation and the data from literatures. It is known that the asymptomatic infection of COVID-19 exists. As reported by Bai *et al*[25], 1 of 6 cases was asymptomatic infected, with the proportion of asymptomatic of 0.17. As reported by Pan *et al*[26], 2 of 3 cases was asymptomatic infected, with the proportion of asymptomatic of 0.67.There was a research showed that the asymptomatic infection would shed SARS-CoV-2 for 5 days[27]. However, there was not enough evidence to provide the clear epidemiological estimates of the parameters *p, κ, ω’*, and *γ’* which were related to asymptomatic characteristics. Although there was clinical evidence to calculate the parameters related to symptomatic cases such as incubation period, fatality rate, and duration from symptoms onset to diagnosis[1, 2], more epidemiological data are needed to explore the parameters. However, we performed a sensitivity analysis of all the nine parameters from the literatures, and we found that the models was slightly sensitive to parameters *p, ω*, and *γ* but not sensitive to parameters *κ, c, ε, f, ω’*, and *γ’*. Therefore, our results might be affected slightly by the parameter estimation.

Since we could not obtain the first-hand data, the results of our findings might have some uncertainties. However, our study aimed to develop and provide an age-specific transmission model for the public health department who has the big data to investigate the age-specific transmissibility in real world scenarios.

## Conclusions

By calculating the published data, our model showed that SARS-CoV-2 has a high transmissibility among adults and elder people but low transmissibility among children and young people. Our results provide a mathematical model for investigating the age-specific transmissibility of SARS-CoV-2. More data and studies are needed to estimate the age-specific transmissibility in real world scenarios to better understand the characteristics of the widely spread virus.

## Data Availability

Not applicable.

## Abbreviations

SARS-CoV-2: severe acute respiratory syndrome coronavirus 2
COVID-19: coronavirus disease 2019
*R*_0_: basic reproduction number
CI: confidence interval
BHRP: Bats-Hosts-Reservoir-People
RP: Reservoir-People
SEIARW: susceptible – exposed – symptomatic – asymptomatic – recovered – reservoir
SAR: secondary attack rate
*R*^2^: coefficient of determination.

## Acknowledgements

Not applicable.

## Availability of data and materials

Not applicable.

## Authors’ contributions

TMC and ZYZ designed research; TMC, YHS, BHZ, and ZYZ conceived the experiments, TMC, ZYZ, YZZ, JWX, ZL, JR, XCL, YW, MY, LL, SSY, JL, RYL, FX, YYS, and YCC conducted the experiments and analyzed the results; TMC, ZYZ, and QQH wrote the manuscript. All authors read and approved the final manuscript.

## Consent for publication

Not applicable.

## Ethics approval and consent to participate

Not applicable.

